# Projecting Introduction Timelines for Cholera Conjugate Vaccine and Typhoid-Cholera Conjugate Combo Vaccine: A Scenario-Based Forecasting Approach

**DOI:** 10.64898/2026.11.09.26367688

**Authors:** Yunjin Yum, Yong-ha Hwang, Wongyeong Choi, Victor Raynaud, Julia Lynch, Jung-Seok Lee

**Affiliations:** International Vaccine Institute, Seoul, Republic of Korea; Gavi, the Vaccine Alliance

## Abstract

**Objective:** This study aimed to project introduction timing and pathways of cholera conjugate vaccine (CCV) and combined typhoid–cholera conjugate vaccines (TCV/CCVs) in 39 low- and middle-income countries and to assess potential barriers to their routine immunization integration.

**Methods:** Accelerated failure time models were fitted using oral cholera vaccine (OCV) introduction as a proxy for CCV adoption. Country-level estimates were standardized using observed post-WHO-prequalification introduction intervals for analogous vaccine transitions. TCV/CCV introduction timing and pathways were projected by combining standardized CCV estimates with a TCV introduction priority index based on typhoid burden and previous vaccination experience. Potential barriers to routine immunization integration were assessed using semi-quantitative scoring approach.

**Results:** Gavi, the Vaccine Alliance (Gavi) eligibility, greater previous OCV experience, and higher cholera intensity were associated with earlier introduction times. After standardization, the median projected time to CCV introduction was 6.9 years after WHO prequalification, with 12 countries introducing within five years. The TCV/CCV projections comprised three pathways: CCV introduction followed by transition to TCV/CCV, direct TCV/CCV introduction and continued CCV use without TCV/CCV introduction during the analysis period. Projected introduction timing was not consistently aligned with routine integration barrier levels for either vaccine.

**Conclusions:** These scenario-based projections provide an early indication of where and when demand may emerge and which countries may require additional programmatic or financing support. The findings can inform demand forecasting, market assessment, manufacturing and supply planning, investment decisions, and introduction preparedness, but the projections should be recalibrated as product-specific evidence becomes available.

## Introduction

Cholera remains an important public health challenge in low- and middle-income countries (LMICs), particularly where access to safe water, sanitation, and hygiene is limited and health systems are affected by conflict, population displacement, or climate-related emergencies (1, 2). An estimated 1.3–4.0 million cases and 21,000–143,000 associated deaths occur globally each year (2). While improvements in water and sanitation provide the most sustainable means of preventing cholera, these require substantial long-term investment. Vaccination therefore remains an important component of integrated cholera control alongside surveillance, community engagement, and water and sanitation interventions (2-4).

Oral cholera vaccines (OCVs) are currently used in preventive and outbreak-response campaigns in populations at risk (4, 5). However, protection may decline over time, vaccine efficacy is lower among young children than among older children and adults, and revaccination may be required to maintain protection (4, 6). Periodic global supply constraints have also restricted preventive vaccination, with preventive campaigns suspended in 2022 and resumed only after vaccine supply improved in 2026 (4, 7). These challenges highlight the need for complementary vaccines that may provide more durable protection, particularly in younger children, and support sustained vaccination strategies.

Cholera conjugate vaccines (CCVs) are being developed to overcome several of these limitations. Conjugation of the *Vibrio cholerae* O-specific polysaccharide (OSP) to a carrier protein is intended to generate a T-cell-dependent immune response and immunological memory. Preclinical findings have supported further development of an OSP tetanus toxin heavy chain fragment (rTTHc) CCV candidate, which is currently undergoing clinical evaluation in adults and children (8, 9). If its safety, efficacy, and duration of protection are established, CCV may offer opportunities for delivery through routine immunization or other sustained preventive strategies, thereby expanding the range of available approaches to cholera prevention and control.

Typhoid fever also imposes a substantial burden in many LMICs and shares several underlying risk factors with cholera, including inadequate access to safe water and sanitation. Typhoid conjugate vaccines (TCVs) are available and recommended for use in typhoid-endemic settings (10). Where the burden, target populations, and vaccination strategies for the two diseases align, a combined typhoid–cholera conjugate vaccines (TCV/CCVs), currently under development, could simplify programme implementation, reduce delivery contacts, and expand opportunities to protect children against both diseases (11). However, its potential value and likelihood of introduction may differ across countries according to cholera and typhoid burden, national policy priorities, and previous OCV and TCV use.

Development and sustainable supply of CCV and TCV/CCV will require investment in clinical development, regulatory approval, manufacturing capacity, and supply preparation. Public health need alone does not necessarily translate into vaccine demand because national adoption also depends on product availability, financing, policy priorities, delivery capacity, and previous vaccination experience. Demand forecasting is therefore an important component of the business case for vaccines under development. Such forecasts estimate which countries may introduce a vaccine, when introduction may occur, and how many doses may subsequently be required, thereby informing assessments of potential market size, manufacturing needs, and investment viability, including expected net present value (NPV) and return on investment (ROI) (12, 13). Among these inputs, country introduction timing is particularly important because it determines when vaccine demand begins and how it develops over time. For CCV and TCV/CCV, forecasting must also account for alternative product pathways, including initial CCV introduction followed by transition to TCV/CCV, direct introduction of TCV/CCV, or continued use of CCV or TCV alone within the analysis period. Because neither product is currently available for country introduction, directly observed adoption data are unavailable. This study therefore aimed to forecast country-specific introduction years and product-transition pathways for CCV and TCV/CCV in 39 selected LMICs.

## Methods

The study included 39 countries: 37 countries that had submitted at least one request to the global OCV stockpile between 2013, when stockpile was established, and 2025 (14), as well as India and Afghanistan. Although India and Afghanistan were not represented in the stockpile dataset, they were included because both countries were relevant to future CCV introduction. India may consider routine use of cholera vaccination, for which OCV has not been well suited, whereas Afghanistan was included because of its substantial recent acute watery diarrhea (AWD) burden, with 43,292 cases reported between January and May 2026—the highest cumulative total among the countries included in the corresponding World Health Organization (WHO) epidemiological update (15).

Country-specific introduction timing and product-transition pathways for CCV and TCV/CCV were predicted through three sequential steps. First, CCV introduction times were estimated using an accelerated failure time (AFT) model fitted to historical OCV introduction data with relevant covariates. Second, the model-based estimates were calibrated by standardizing them to empirically observed post--WHO-prequalification (WHO PQ) introduction intervals for analogous vaccine transitions. Third, for projecting TCV/CCV introduction timing and product-transition pathways, TCV introduction priority index and previous country experience with OCV and TCV were incorporated in the calibrated country-specific CCV introduction year baseline. Following these three forecasting steps, a semi-quantitative scoring system was developed to assess countries according to potential barriers to integrating each of CCV and TCV/CCV into Expanded Programme on Immunization (EPI). The overall modeling frameworks are depicted in Figure S1.

### CCV introduction years

#### Accelerated failure time (AFT) model

To estimate the time to CCV introduction, parametric survival analysis was conducted using AFT models. The AFT approach was selected because it directly models the association between country-level characteristics and the timing of vaccine introduction (16). Within this framework, covariates are assumed to accelerate or decelerate the time to introduction, facilitating the interpretation of factors associated with earlier or later vaccine introduction.

The general form of the AFT model is (16, 17):

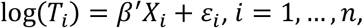

where *T*_*i*_ denotes the time to vaccine introduction for country *i, X*_*i*_ is a *p* × 1 vector of observed covariates, *p* denotes the number of covariates, *β* is the corresponding *p*-dimensional vector of regression coefficient, and *ε* _*i*_ represents the error term. Four parametric distributions were considered: exponential, Weibull, log-logistic, and log-normal distributions (18, 19).

#### Dependent variable

Because CCV is currently in development, OCV introduction data were used as a proxy for future CCV introduction. Accordingly, the dependent variable was defined as the time from the WHO pre-qualification (PQ) of Shanchol in September 2011 (index date) to the first OCV introduction. However, because the global OCV stockpile was established in 2013, stockpile data were unavailable between WHO PQ of Shanchol (20) and stockpile establishment. To address this gap, information on OCV use during this period was further identified through a literature review (21) and included in the outcome definition if it involved a reactive campaign followed by an additional OCV stockpile use. Countries were followed from the index date until either vaccine introduction or the end of the study period (2025). Countries that had not yet introduced the vaccine by the end of the observation period were treated as right-censored. The observation period was defined through 2025 because OCV stockpile data were available and updated through that year at the time of the current study (22).

#### Independent variables

An initial set of independent variables was identified based on previous literature (19, 23) and organized into three domains: health-related factors, socio-economic factors, and modifying factors. The variable type, time-varying status, and data source for each variable are presented in Table 1.

**Table 1.** Independent variables.

| Independent variables | Type | Time-varying | Data source |
|---|---|---|---|
| ***Health*** |  |  |  |
| Natural log DALYs | Continuous | Yes | IHME GBD (24) |
| MCV coverage (%) | Continuous | Yes | WHO Immunization data (25) |
| DTP3 coverage (%) | Continuous | Yes | WHO Immunization data (25) |
| PCV introduction status | Binary | Yes | VIEW-hub (26) |
| OCV experience | Continuous | No | GTFCC (22) |
| Cholera intensity | Binary | Yes | WHO weekly report/Cholera cases & deaths (Official annual data reported to WHO) (27) |
| ***Socio-economic*** |  |  |  |
| Natural log GNI per capita | Continuous | Yes | World Bank (28) |
| Natural log population | Continuous | Yes | UN population division (29) |
| Region (Geopolitical region) |  |  | World Bank (28) |
| East Asia & Pacific | Binary | No |  |
| Latin America & Caribbean | Binary | No |  |
| Middle East, North Africa, Afghanistan & Pakistan | Binary | No |  |
| South Asia | Binary | No |  |
| Sub-Saharan Africa | Binary | No |  |
| Democracy score | Continuous | Yes | World Bank (30) |
| Natural log cost per hospital bed-day | Continuous | Yes | WHO CHOICE (31) |
| ***Modifying factors*** |  |  |  |
| Neighboring introducing countries | Continuous | Yes | GTFCC (22) |
| WHO position paper | Binary | Yes | WHO (5) |
| Gavi-eligibility | Binary | Yes | Gavi (32)/World Bank (28) |

DALYs, disability-adjusted for life year; MCV, measles-containing vaccine; DTP3, Diphtheria tetanus toxoid and pertussis; PCV, pneumococcal conjugate vaccine; OCV, oral cholera vaccine; GNI, gross national income; Gavi, Gavi, the Vaccine Alliance; IHME GBD, Institute for Health Metrics and Evaluation Global Burden of Disease; GTFCC, The Global Task Force on Cholera Control (GTFCC); WHO CHOICE, World Health Organization CHOosing Interventions that are Cost-Effective.

In the health domain, higher cholera burden was hypothesized to increase the perceived public health benefits of vaccination and therefore accelerate vaccine introduction. Diphtheria tetanus toxoid and pertussis and MCV coverage were included as proxies for immunization program capacity (33). Previous introduction of pneumococcal conjugate vaccine (PCV) was considered an indicator of a country’s propensity to introduce new vaccines; countries that had introduced these vaccines earlier were hypothesized to introduce CCV sooner than countries with delayed introduction. OCV experience was measured using a weighted frequency of previous OCV requests, which accounted for both the number and context of OCV deployments (i.e., hotspot response, outbreak response, or humanitarian crisis) (22). Because this measure spans the entire observation window, it includes requests occurring after a country’s first OCV introduction and is therefore intended to capture each country’s overall accumulated engagement with OCV as of the forecasting origin rather than a temporally antecedent predictor of its own first introduction. Cholera intensity was defined based on whether cholera cases or deaths were reported each year, using official annual data reported to WHO through 2024 (27) and WHO weekly reports for 2025. For WHO weekly reports, countries were classified as having high cholera intensity if their outbreak risk levels were consistently reported as acute crisis or active in all reports published during the year.

In the socio-economic domain, gross national income (GNI) per capita was included to account for differences in economic capacity that could influence vaccine introduction decisions. Democracy score was used to capture differences in governance and decision-making environments, with the hypothesis that more democratic countries may introduce new vaccines more rapidly. Cost per hospital bed-day was included as a proxy for the economic consequences, under the assumption that countries facing higher treatment costs may perceive greater value from vaccination.

In the context of modifying factors, the number of neighboring introducers was included to capture potential policy diffusion effects, whereby vaccine introduction decisions in neighboring countries may influence national decision-making. WHO position paper was included to assess whether global recommendations accelerated introduction decisions (5). Gavi, the Vaccine Alliance (Gavi) eligibility, whether a country meets the criteria for Gavi support, was included as a country-level predictor of vaccine introduction timing.

Missing values were addressed using variable-specific imputation methods, including United Nations (UN) Population Division projections, Gross domestic product (GDP) deflator adjustment, and last observation carried forward (LOCF).

#### Model selection and input data

Model selection was based on the Akaike Information Criterion (AIC), Bayesian Information Criterion (BIC), and the epidemiological plausibility of the estimated associations between covariates and the outcome. The final selected AFT model was applied to country-specific covariate profiles projected to the expected WHO PQ year of CCV (2034) to estimate the timing of CCV introduction. Future covariate values were derived using variable-specific methods; specifically, Gavi eligibility in 2034 was estimated based on projected GNI per capita. The data used to estimate country-specific CCV introduction years are presented in Table S1 and rationale for estimated WHO PQ year of CCV is presented in Table S2.

### Standardization

Because CCV represents a conjugate-vaccine transition from the already available OCV rather than the introduction of a vaccine targeting an entirely new disease, it was expected to be introduced more rapidly than a fully new vaccine. To derive an empirically plausible adoption window, we examined country-level introduction intervals following WHO PQ for analogous vaccine replacements or transitions, including PCV, rotavirus vaccines (RV), meningococcal vaccines, and polio vaccines. The transitions examined included PCV7 to PCV10 or PCV13, PCV10 to PCV13, RV1 to RV5, meningococcal group A polysaccharide vaccine to meningococcal group A conjugate vaccine (MenAfriVac), and the replacement of trivalent oral poliovirus vaccine (OPV) with bivalent tOPV with bOPV accompanied by the introduction of inactivated poliovirus vaccine (IPV) (26, 34, 35). Across the eligible country-level transition observations, the interval between WHO PQ and introduction ranged from 1 to 12 years. The AFT projected CCV introduction times were therefore rescaled to this observed range using the min–max standardization method (36), preserving the relative ordering of countries while improving the empirical plausibility of the projected introduction timing.

### TCV/CCV introduction years - Typhoid introduction priority index

A semi-quantitative scoring system was used to assess each country’s priority for TCV introduction. The scoring system comprised two indicators: typhoid disease burden and previous typhoid vaccination experience. Disease burden scores were assigned according to quintiles of the country’s mean annual typhoid DALY rates from 2019 to 2023. Vaccination experience scores were based on the introduction strategy (i.e., national immunization programme (NIP) or campaign) and year of introduction. The two indicator scores were summed and categorized to derive the TCV introduction priority index, which was subsequently used to determine country-specific adjustments to TCV/CCV introduction and CCV-to-TCV/CCV transition timing. Detailed scoring criteria and timing adjustments are provided in Tables S3.

If the projected CCV introduction preceded assumed TCV/CCV WHO PQ (2036) (Table S2), countries were assumed to introduce CCV first and subsequently transition to TCV/CCV. Among countries expected to introduce CCV after TCV/CCV WHO PQ, those with high TCV introduction priority and prior experience with both OCV and TCV were assumed to introduce TCV/CCV earlier than the CCV baseline, whereas those with high priority but limited vaccine experience were assigned a similar timeline to CCV introduction. Countries with low-to-moderate priority were assumed to introduce TCV/CCV later than the CCV baseline. In all three cases, CCV was replaced by TCV/CCV. Countries with lower priority were assumed not to introduce TCV/CCV within the analysis period.

### EPI barrier

To assess countries according to potential barriers to EPI integration for each of CCV and TCV/CCV, a semi-quantitative scoring system was developed. Country-level indicators were selected based on the WHO vaccine introduction decision framework (37, 38) and existing evidence (23) and grouped into three domains: disease characteristics, immunization system capacity, and country readiness. These indicators included disease intensity, previous OCV stockpile requests, experience with newer vaccine introductions, MCV coverage, political will, and Gavi eligibility. Each indicator was standardized and scored, with lower scores indicating fewer barriers. Indicators were weighted at 10% for disease characteristics, 30% for immunization system capacity, and 10% for country readiness, reflecting the greater importance of immunization system capacity for EPI integration. Weighted scores were summed and divided into tertiles to classify countries into low-, medium-, or high-barrier groups. For the CCV assessment, all indicators except typhoid disease intensity were included, whereas the TCV/CCV assessment included all indicators. Detailed indicator definitions and scoring criteria are provided in Table 2.

**Table 2.**
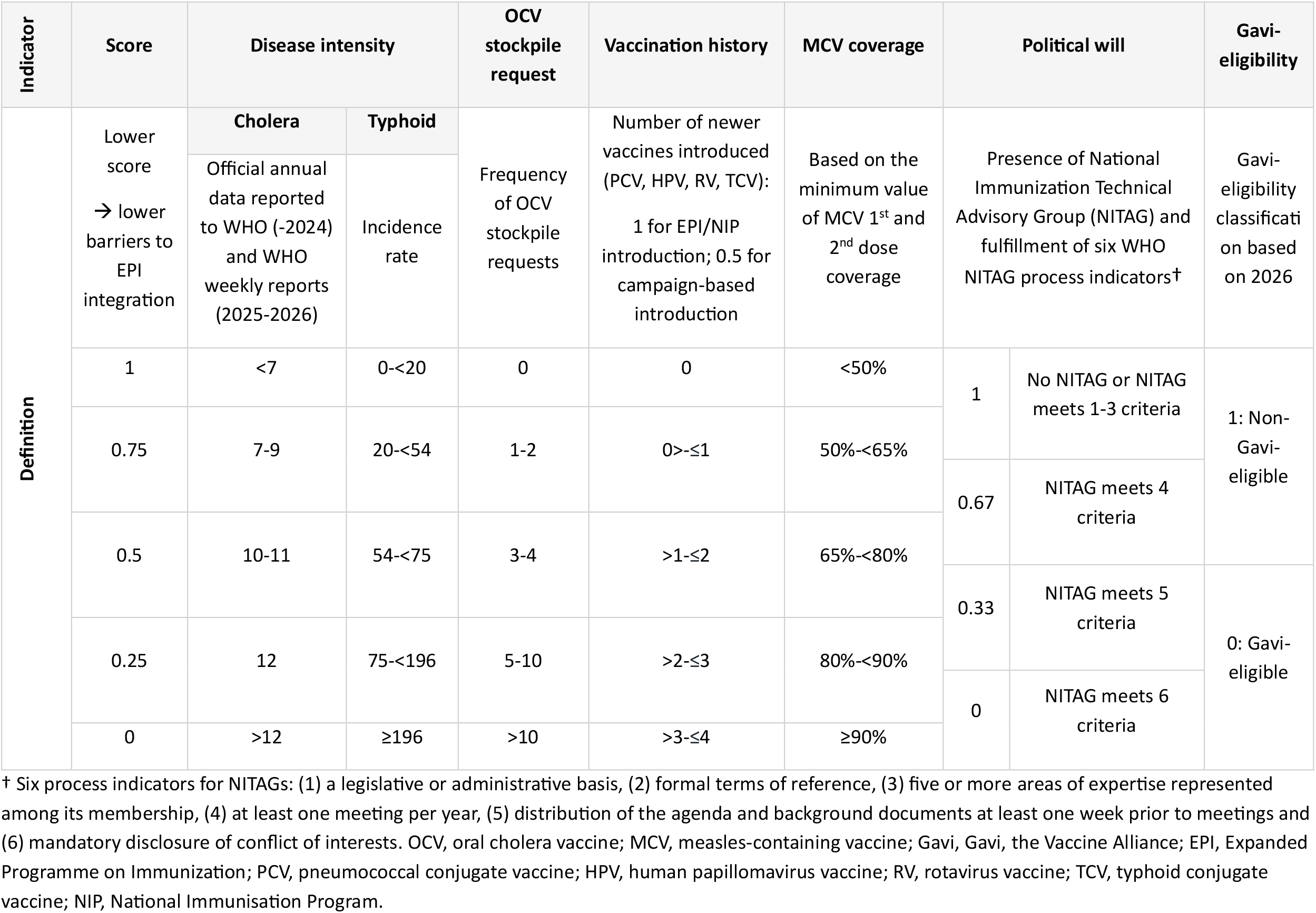
Scoring of the EPI barrier.

| Indicator | Score | Disease intensity |  | OCV stockpile request | Vaccination history | MCV coverage | Political will |  | Gavi-eligibility |
|---|---|---|---|---|---|---|---|---|---|
| Definition | Lower score<br>→ lower barriers to EPI integration | **Cholera**<br>Official annual data reported to WHO (-2024) and WHO weekly reports (2025-2026) | **Typhoid**<br>Incidence rate | Frequency of OCV stockpile requests | Number of newer vaccines introduced (PCV, HPV, RV, TCV):<br>1 for EPI/NIP introduction; 0.5 for campaign-based introduction | Based on the minimum value of MCV 1<sup>st</sup> and 2<sup>nd</sup> dose coverage | Presence of National Immunization Technical Advisory Group (NITAG) and fulfillment of six WHO NITAG process indicators† |  | Gavi-eligibility classification based on 2026 |
|  | 1 | <7 | 0-<20 | 0 | 0 | <50% | 1 | No NITAG or NITAG meets 1-3 criteria | 1: Non-Gavi-eligible |
|  | 0.75 | 7-9 | 20-<54 | 1-2 | 0>-≤1 | 50%-<65% | 0.67 | NITAG meets 4 criteria |  |
|  | 0.5 | 10-11 | 54-<75 | 3-4 | >1-≤2 | 65%-<80% | 0.33 | NITAG meets 5 criteria | 0: Gavi-eligible |
|  | 0.25 | 12 | 75-<196 | 5-10 | >2-≤3 | 80%-<90% | 0 | NITAG meets 6 criteria |  |
|  | 0 | >12 | ≥196 | >10 | >3-≤4 | ≥90% |  |  |  |

† Six process indicators for NITAGs: (1) a legislative or administrative basis, (2) formal terms of reference, (3) five or more areas of expertise represented among its membership, (4) at least one meeting per year, (5) distribution of the agenda and background documents at least one week prior to meetings and (6) mandatory disclosure of conflict of interests. OCV, oral cholera vaccine; MCV, measles-containing vaccine; Gavi, Gavi, the Vaccine Alliance; EPI, Expanded Programme on Immunization; PCV, pneumococcal conjugate vaccine; HPV, human papillomavirus vaccine; RV, rotavirus vaccine; TCV, typhoid conjugate vaccine; NIP, National Immunisation Program.

## Results

Between 2011 and 2025, 34 of the 39 study countries had introduced OCV, while the remaining five countries were treated as right-censored: Philippines, Burundi, and Iran, had submitted requests to the OCV stockpile but had not received approval, whereas India and Afghanistan had not submitted requests to the stockpile at all. From the WHO PQ of OCV in September 2011, the observed time to OCV introduction ranged from approximately 7 months to 14 years (please see Figure S2).

The three candidate models were selected from the initial specifications based on AIC, BIC, statistical significance, and epidemiological plausibility. Among the three candidate models presented in Table 3, Model 3 with Weibull distribution was selected as the final model, as it showed the lowest AIC and BIC alongside significant p-values. In Model 3, Gavi eligibility was associated with a 22% shorter time to vaccine introduction than non-eligibility (0.78; 95% CI, 0.61–0.98). Greater OCV experience and higher cholera intensity were also associated with shorter times to introduction, by 18% (0.82; 95% CI, 0.78– 0.86) and 27% (0.73; 95% CI, 0.56–0.94), respectively.

**Table 3.** Analysis results of the Weibull AFT models.

| Model | Variable | Proportionate change | 95% CI Lower | 95% CI Upper | *p* | AIC | BIC |
|---|---|---|---|---|---|---|---|
| Model 1 | GNI per capita | 0.84 | 0.67 | 1.06 | 0.14 | 50.15 | 73.22 |
|  | Gavi-eligibility | 0.62 | 0.43 | 0.91 | 0.01 |  |  |
|  | OCV experience | 0.82 | 0.78 | 0.86 | 0.00 |  |  |
|  | Cholera intensity | 0.69 | 0.52 | 0.91 | 0.01 |  |  |
|  | Constant | 81.14 | 11.93 | 551.59 | 0.00 |  |  |
| Model 2 | Cholera burden (DALYs) | 0.94 | 0.84 | 1.04 | 0.23 | 51.04 | 74.10 |
|  | Gavi-eligibility | 0.78 | 0.62 | 0.97 | 0.03 |  |  |
|  | OCV experience | 0.83 | 0.79 | 0.88 | 0.00 |  |  |
|  | Cholera intensity | 0.76 | 0.59 | 0.97 | 0.03 |  |  |
|  | Constant | 26.16 | 15.35 | 44.57 | 0.00 |  |  |
| Model 3 | Gavi-eligibility | 0.78 | 0.61 | 0.98 | 0.03 | 50.46 | 69.68 |
|  | OCV experience | 0.82 | 0.78 | 0.86 | 0.00 |  |  |
|  | Cholera intensity | 0.73 | 0.56 | 0.94 | 0.02 |  |  |
|  | Constant | 20.10 | 15.04 | 26.85 | 0.00 |  |  |

AIC, Akaike Information Criterion; BIC, Bayesian Information Criterion; GNI, gross national income; Gavi, Gavi, the Vaccine Alliance; OCV, oral cholera vaccine; DALY, disability-adjusted life year.

Kaplan-Meier curves for both CCV introductions before and after standardization are presented in Figure 1. Before standardizing, Model 3 estimated that 50% of the target countries would introduce CCV within 10.7 years (Figure 1 (A)). After rescaling, the median projected time to introduction decreased to 6.9 years (Figure 1 (B)). Twelve countries—South Sudan, Democratic Republic of the Congo, Sudan, Ethiopia, Mozambique, Malawi, Nigeria, Zambia, Yemen, Pakistan, Haiti, and Somalia—were projected to introduce CCV within 5 years (Figure 2). Most of these countries were classified as having acute or active cholera risk in the 2026 WHO report. In contrast, the eight countries projected to require at least 10 years—Lebanon, Guinea, Iraq, Côte d’Ivoire, Dominican Republic, Lao People’s Democratic Republic, Iran, and the Philippines—had no or only one previous OCV deployment and relatively few cholera cases reported to WHO. Countries with stronger immunization systems and higher cholera intensity generally had low barriers to CCV integration into EPI. However, projected introduction timing did not consistently align with barrier levels, as some countries expected to introduce CCV early still faced substantial implementation constraints. (Figure 2, Figure S3 (A)). For example, Sudan and Yemen were projected to introduce CCV relatively early but was classified as having high barriers to routine immunization integration. This would be due in part to limited experience introducing newer vaccines, and relatively low MCV coverage.

**Figure 1.**
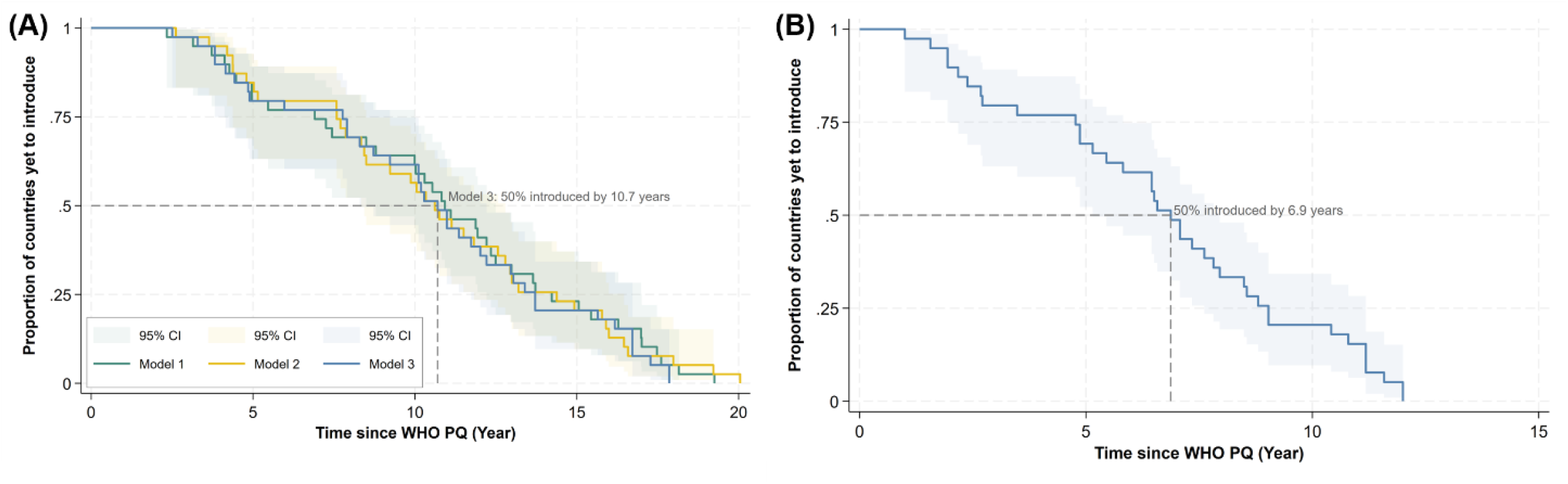
Time to CCV introduction: (A) before rescaling for Model 1-3 (B) after rescaling Model 3.

**Figure 2.**
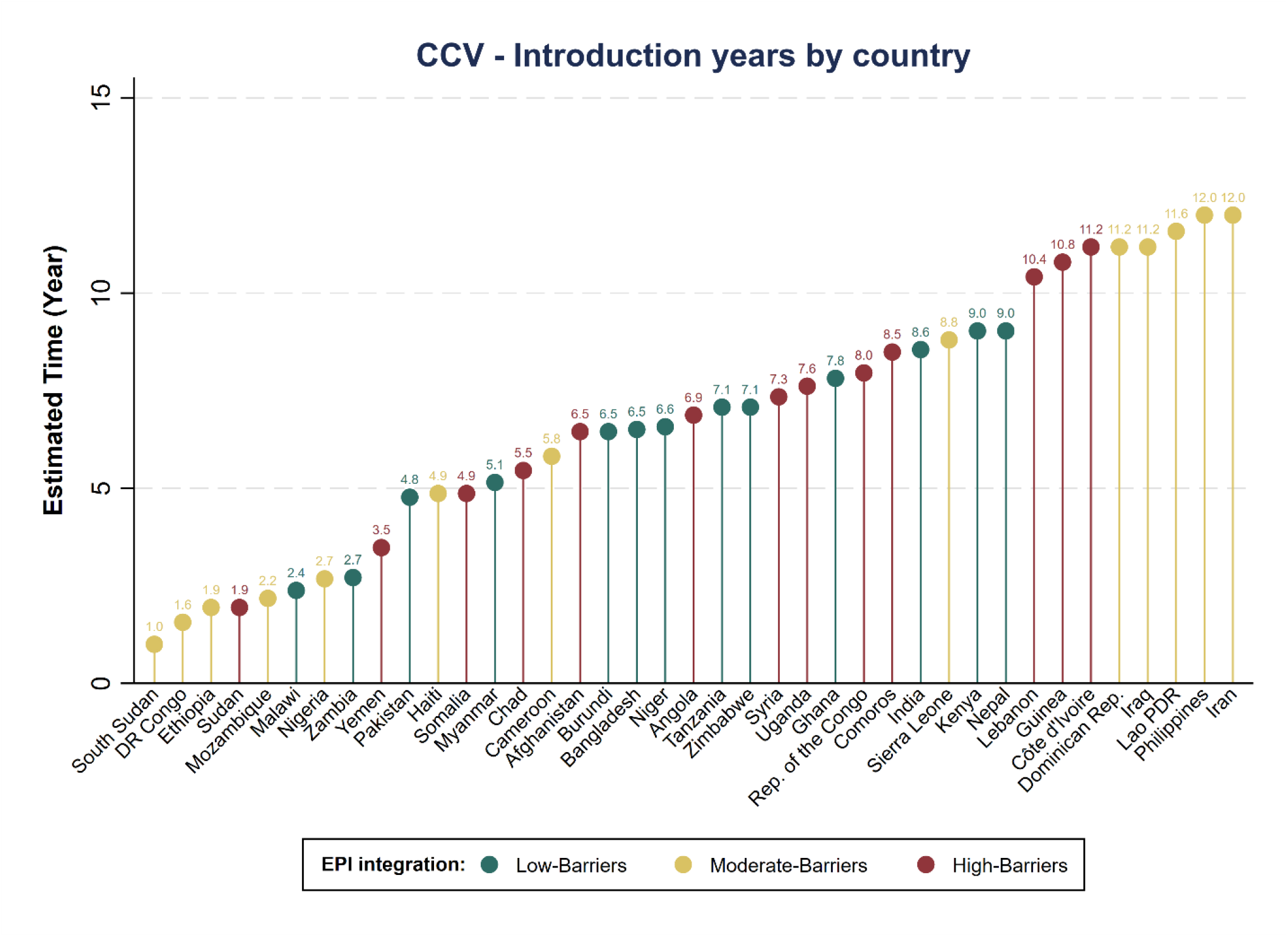
CCV introduction years by country with EPI barriers. EPI, Expanded Programme on Immunization; DR Congo, Democratic Republic of the Congo; Rep. of the Congo, Republic of the Congo; Dominican Rep., Dominican Republic; Lao PDR, Lao People’s Democratic Republic.

Projected TCV/CCV introduction timing and vaccine-transition pathways are presented in Figure 3. Eight countries—South Sudan, Democratic Republic of the Congo, Sudan, Ethiopia, Mozambique, Malawi, Nigeria, and Zambia—were projected to introduce CCV in 2035–2036, and subsequently transition to TCV/CCV between 2038 and 2040. Countries with baseline CCV introduction projected after TCV/CCV WHO PQ and with high or low-moderate TCV introduction priority were assumed to skip CCV and introduce TCV/CCV directly, with the timing shifted 1 year earlier, left unchanged, or shifted 1 year later according to the scoring adjustment. Seven countries with low TCV introduction priority—Syria, Lebanon, Haiti, Dominican Republic, Iraq, Iran, and Lao People’s Democratic Republic—were projected to introduce CCV only and not TCV/CCV within the analysis period.

**Figure 3.**
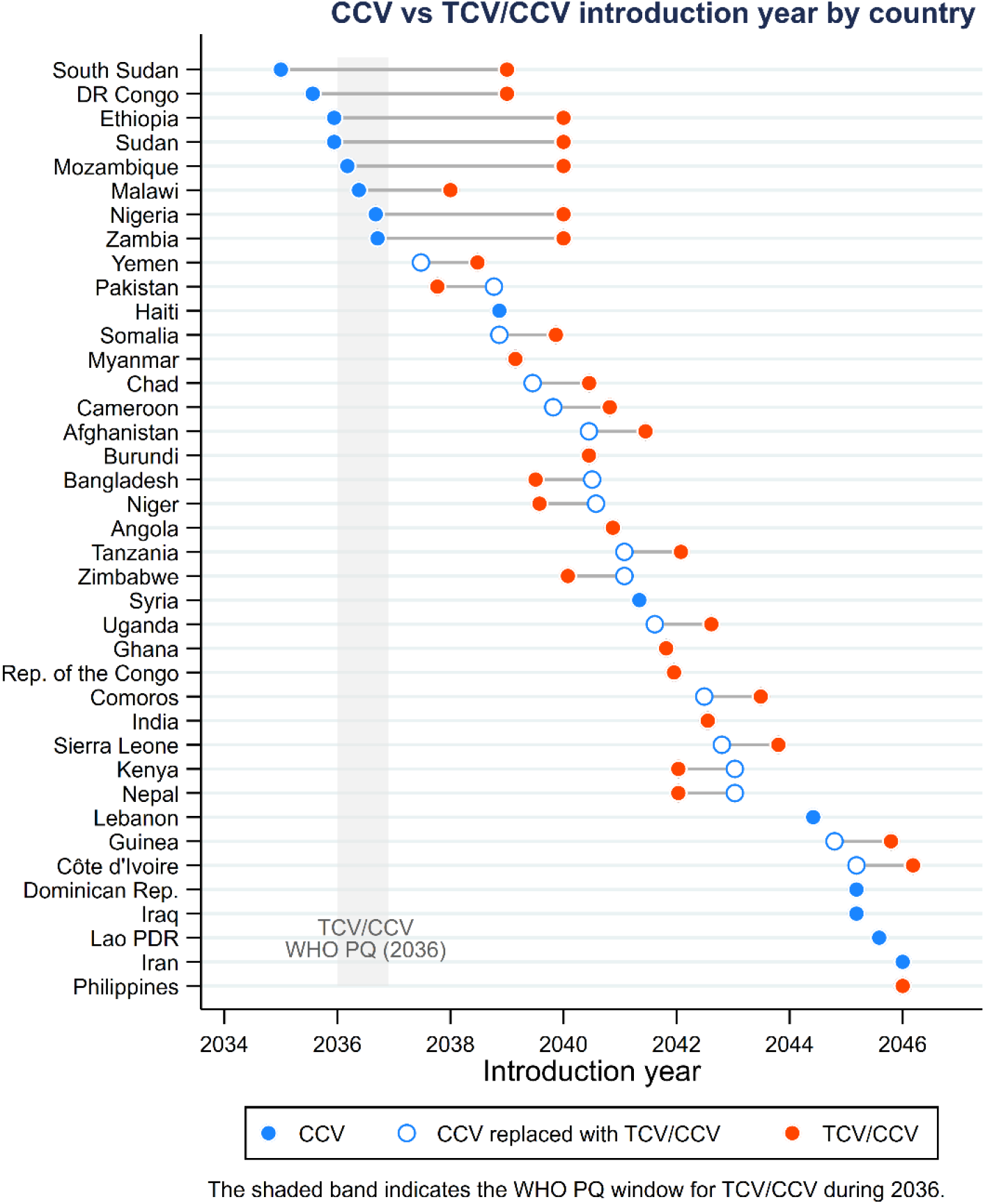
CCV and TCV/CCV introduction year by country and vaccine-transition pathways by country. DR Congo, Democratic Republic of the Congo; Rep. of the Congo, Republic of the Congo; Dominican Rep., Dominican Republic; Lao PDR, Lao People’s Democratic Republic.

Figure 4 presents the projected introduction timing alongside the assessed barriers to integration into EPI for TCV/CCV (Figure S3 (B)). The TCV/CCV assessment included 32 countries, excluding the seven countries projected to introduce CCV only due to lower TCV introduction priority. Yemen was similarly projected to introduce TCV/CCV relatively early but was classified as having high integration barriers because of its limited experience with newer vaccine introductions and low typhoid intensity. In contrast, Ghana, Kenya, Nepal, and Tanzania were classified as having relatively low integration barriers despite their comparatively long projected delays in introduction, owing to their Gavi eligibility and experience with newer vaccine introductions.

**Figure 4.**
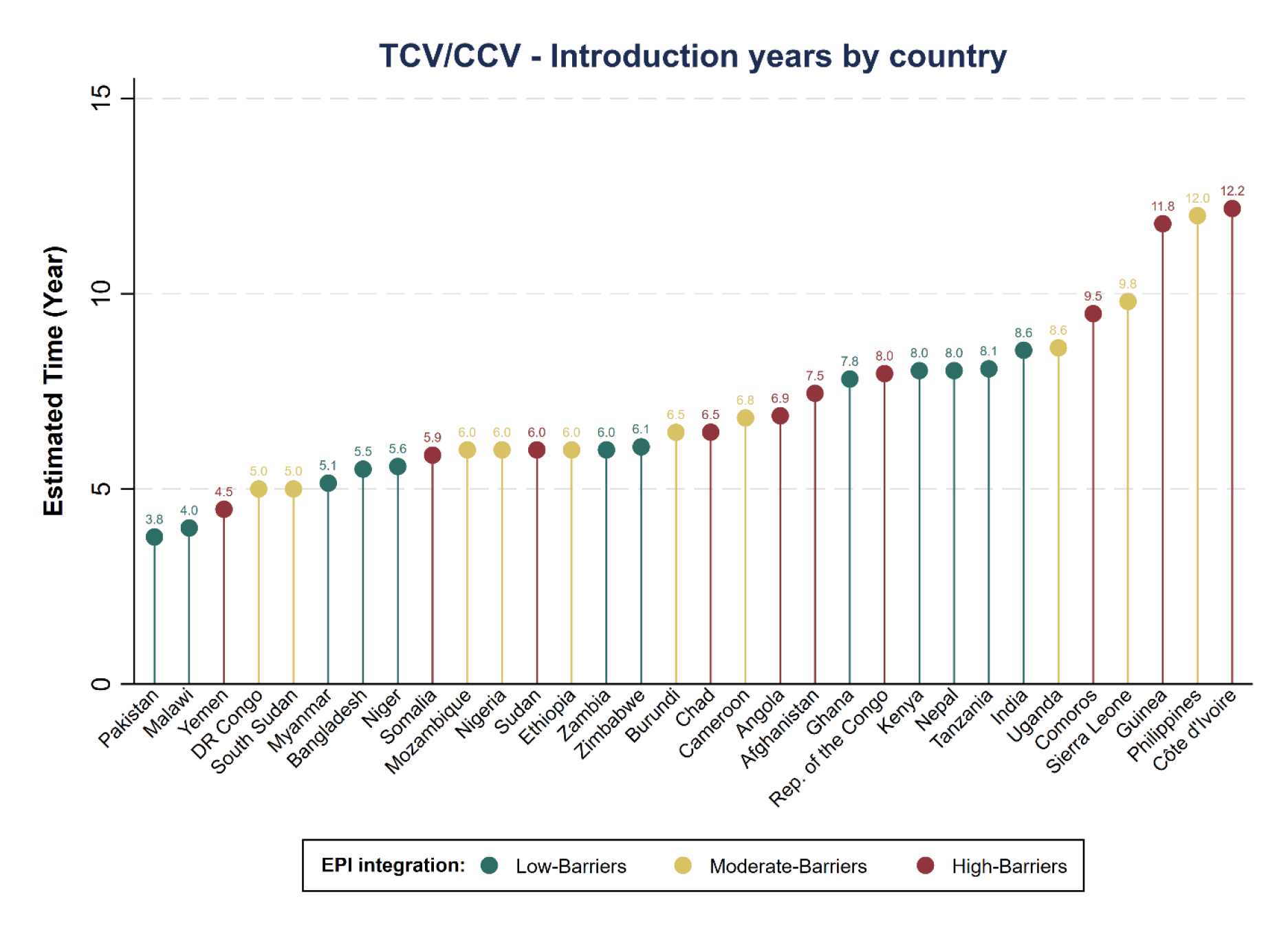
TCV/CCV introduction years by country with EPI barriers. EPI, Expanded Programme on Immunization; DR Congo, Democratic Republic of the Congo; Rep. of the Congo, Republic of the Congo; Dominican Rep., Dominican Republic; Lao PDR, Lao People’s Democratic Republic.

## Discussion

This study estimated country-specific timelines for CCV and TCV/CCV introduction and assessed barriers to their integration into EPI. Gavi eligibility, previous OCV experience, and cholera intensity were associated with the timing of CCV introduction. Gavi eligibility may reduce financial constraints and facilitate access to procurement, planning, and technical support. Previous OCV experience may indicate operational capacity and sustained policy interest in cholera control, while greater cholera intensity may increase the urgency of vaccination. Earlier projected introduction generally corresponded to lower barriers to EPI integration. A similar pattern was observed in the TCV/CCV scenarios, with earlier introduction generally corresponding to lower integration barriers. Under these scenarios, countries followed one of three pathways: direct introduction of TCV/CCV, transition from CCV to TCV/CCV, or no TCV/CCV introduction within the analysis period.

These findings are broadly consistent with previous research on Haemophilus influenzae type b (Hib) vaccine adoption, which identified disease burden and Gavi support as factors associated with shorter decision times (19). Research on barriers to PCV introduction has similarly highlighted Gavi eligibility as an important facilitator (23). Together, these findings suggest that financial and technical support can help translate public health need into vaccine adoption. Although higher vaccine prices have been associated with delayed vaccine introduction in previous studies, vaccine price was not included because CCV and TCV/CCV are still under development and their prices remain unknown.

It is worth noting that early projected introduction in South Sudan, Sudan, Somalia, Yemen, and the Democratic Republic of the Congo partly reflects their extensive OCV stockpile experience and recent cholera reporting to WHO. However, limited immunization capacity in these countries may pose challenges to early vaccine introduction. The EPI integration barrier analysis provides an indirect indication of the feasibility of routine integration, taking such constraints into account. In addition, the CCV barrier classifications used in our analysis do not necessarily align with countries’ current or expected near-term interest in OCV. For example, Somalia and Sudan showed high interest in OCV but faced high barriers to CCV integration, reflecting low MCV coverage and limited experience introducing newer vaccines. Conversely, some countries with limited apparent interest in OCV faced relatively low integration barriers because of Gavi eligibility and functional NITAGs meeting all six assessed criteria. OCV interest reflects current demand for cholera vaccination, whereas CCV integration barriers reflect the broader capacity and enabling environment for routine delivery in addition to the disease burden intensity. These assessments should therefore be interpreted alongside country-specific epidemiological, programmatic, and policy contexts. Depending on its eventual characteristics and delivery strategy, CCV could support a shift from reactive outbreak response toward more sustained preventive cholera control, with integration into routine immunization requiring sufficient country-level capacity.

This study has several limitations. First, historical OCV introduction patterns were used as a proxy because no real-world CCV introduction data exist. Thus, the proxy reflects both a country’s decision to request the vaccine and the International Coordinating Group’s (ICG) approval rather than the country’s introduction decision alone. In addition, not only future CCV adoption through routine immunization but also, campaign - and outbreak-based use of CCV may differ from the campaign- and outbreak-based use of OCVs. Second, the small number of OCV introduction events limited statistical power and increased the risk of overfitting. Thus, projections may be sensitive to the selected AFT distribution and model specification. Third, future covariate values were estimated using LOCF, historical averages, or linear extrapolation, which may not capture unexpected changes in disease burden, financing, or health-system capacity. Fourth, although Gavi eligibility, previous OCV experience, and cholera intensity were associated with the timing of CCV introduction in the AFT model, these associations should not be interpreted as causal, as these indicators may also reflect unmeasured differences in health-system capacity, financing, and political commitment. In particular, previous OCV experience was measured using weighted request frequency over the entire 2011–2025 observation period and therefore included requests occurring after a country’s first observed vaccine introduction. Its association with the proxy outcome may thus reflect temporal overlap rather than a prospective relationship. This indicator was retained solely to characterize accumulated country engagement with OCV for the scenario-based projections. Fifth, the EPI barrier classifications were based on a semi-quantitative composite score and may not fully capture country-specific implementation constraints. The TCV/CCV timelines also depended on assumed WHO PQ years and predefined transition scenarios rather than an independently estimated statistical model. The estimated introduction years should therefore be interpreted as indicative timelines rather than precise predictions. Finally, it should be noted that the projections in the current study are the authors’ own and do not represent Gavi forecasts, policies, or commitments; any potential support for CCV or TCV/CCV would be considered through Gavi’s Vaccine Investment Strategy process. Even if such support becomes available, countries’ capacity to meet co-financing requirements may affect the feasibility and timing of introduction.

Despite these limitations, this study provides policy- and market-relevant inputs for forecasting future demand for CCV and TCV/CCV. By estimating country-specific introduction timelines, identifying potential product-transition pathways, and assessing barriers to EPI integration, the analysis offers an early indication of where demand may emerge, how rapidly it may develop, and which countries may require additional programmatic or financing support before introduction. These projections may help vaccine developers, manufacturers, commercialization partners, and global health stakeholders assess potential market size, plan manufacturing and supply strategies, and evaluate expected returns on investment under alternative introduction scenarios. At the same time, the findings can support country-level and global planning by highlighting settings where early engagement, evidence generation, financing preparation, and immunization-system strengthening may be needed to facilitate future uptake. As CCV and TCV/CCV candidates progress through clinical development and more product-specific data become available, the forecasting framework should be refined and recalibrated to support more precise demand estimates and more actionable introduction planning.

## Data Availability

The datasets generated and/or analyzed during the current study are available from the GTFCC OCV database

https://apps.epicentremsf.org/public/app/gtfcc.

## Competing interests

We would like to mention that IVI’s involved in current CCV development.

## Acknowledgements

The current study was supported by a grant from Prof. Joon Haeng Rhee through the Korea Support Committee (KSC) for International Vaccine Institute (IVI) (grant number: KSC202601-TCV-CCV). Additionally, the International Vaccine Institute gratefully acknowledges the partnership and financial contributions provided by the governments of the Republic of Korea, Sweden, Austria, Finland, India, Thailand, and the Philippines.

## Notes

### Competing Interest Statement

We would like to mention that IVI has involved in current CCV development.

### Author Declarations

IHME GBD data :https://vizhub.healthdata.org/gbd-results/

