## Supplementary materials for "Projecting Introduction Timelines for Cholera Conjugate Vaccine and Typhoid-Cholera Conjugate Combo Vaccine: A Scenario-Based Forecasting Approach"

### Supplementary Table S1. Input data.

| Independent variables | Methods |
|---|---|
| Population | UN Population Division projections applied. |
| Geopolitical region | Treated as constant (time-invariant variable). |
| PCV | LOCF applied. |
| MCV coverage | Latest 3-year average applied. |
| DTP3 coverage | Latest 3-year average applied. |
| Democracy score | LOCF applied. |
| OCV experience | Treated as constant (time-invariant variable). |
| Cholera intensity | WHO weekly report |
| GNI per capita | GDP deflator adjustment with linear prediction applied. |
| Cost per bed-day | GDP deflator adjustment with linear prediction applied. |
| DALYs | LOCF applied. |
| Neighboring introducers | LOCF applied. |
| WHO position paper | LOCF applied. |
| Gavi eligibility | Estimated based on projected GNI per capita values and the Gavi eligibility threshold. |

PCV, pneumococcal conjugate vaccine; MCV, measles-containing vaccine; DTP3, Diphtheria tetanus toxoid and pertussis; OCV, oral cholera vaccine; GNI, gross national income; DALYs, disability-adjusted for life year; Gavi, Global alliance for vaccines and immunization; LOCF, last observation carried forward; GDP, gross domestic product.

**Supplementary Table S2. Rationale for estimated WHO prequalification (PQ) years.**

| Vaccine | Estimated WHO PQ | Rationale |
|---|---|---|
| **CCV** | 2034 | Based on completion of the ongoing Phase II study in 2027, followed by a pivotal Phase III immunogenicity and safety non-inferiority trial against an OCV comparator (2028–2031), national regulatory approval and WHO PQ preparation (2032–2033), with WHO PQ under the minimum profile (target age ≥1 year) expected in 2034. |
| **TCV/CCV** | 2036 | Based on an assumed first-in-human study in 2028, applying the approximately seven-year development-to-PQ timeline observed for MenFive[1] (2035), and allowing an additional year for combination-vaccine development (2036). |

[1]MenFive is a pentavalent meningococcal conjugate vaccine targeting serogroups A, C, W, Y, and X.

**Supplementary Table S3. Scoring of the TCV introduction priority index and corresponding timing adjustments.**

Lower component scores indicate greater priority for TCV introduction. The summed score was categorized into the TCV introduction priority index, for which a higher index indicates greater priority.

### Table S3-A. Component scoring and derivation of the TCV introduction priority index

| Indicator | Definition | Component score | Scoring criterion | TCV Introduction Priority Index | Total score |
|---|---|---|---|---|---|
| **Typhoid disease burden** | Mean annual typhoid DALYs (2019–2023), categorized into quintiles | 1.00 | 0–<11 | **4** | ≤1.25 |
| | | 0.75 | 11–<66 | | |
| | | 0.50 | 66–<97 | **3** | >1.25–≤1.50 |
| | | 0.25 | 97–<156 | | |
| | | 0.00 | ≥156 | **2** | >1.50–≤1.75 |
| **Typhoid vaccination experience** | Based on vaccination strategy (NIP vs. campaign) and year of introduction | 1.00 | Not yet introduced | | |
| | | 0.75 | Campaign | **1** | >1.75–≤2.00 |
| | | 0.50 | NIP introduction in 2025–2026 | | |
| | | 0.25 | NIP introduction in 2022–2023 | Index derived from the summed component score | |
| | | 0.00 | NIP introduction in 2019–2021 | | |

### Table S3-B. Country-specific adjustment of TCV/CCV introduction and CCV to TCV/CCV transition timing

| TCV priority index | Additional criterion | CCV to TCV/CCV transition adjustment (years)[*] | Direct TCV/CCV introduction adjustment (years)[**] | Interpretation |
|---|---|---|---|---|
| **4** | Prior introduction of both OCV and TCV | **+2** | **−1** | Earlier TCV/CCV adoption relative to other countries |
| **4** | All other countries | **+3** | **0** | No adjustment to direct introduction; transition occurs three years after the WHO PQ |
| **3 or 2** | None | **+4** | **+1** | Later TCV/CCV adoption relative to the CCV baseline |
| **1** | None | **CCV only[***]** | **CCV only[***]** | No TCV/CCV introduction within the analysis period |

[*] Countries projected to introduce CCV prior to the assumed TCV/CCV WHO PQ: South Sudan, Democratic Republic of the Congo, Ethiopia, Sudan, Mozambique, Malawi, Nigeria, and Zambia

[**] Countries expected to introduce CCV after TCV/CCV WHO PQ: Yemen, Pakistan, Somalia, Myanmar, Chad, Cameroon, Afghanistan, Burundi, Bangladesh, Niger, Angola, Tanzania, Zimbabwe, Uganda, Ghana, Republic of the Congo, Comoros, India, Sierra Leone, Kenya, Nepal, Guinea, Côte d'Ivoire, and Philippines

[***] Countries assigned to the CCV-only pathway were assumed not to introduce TCV/CCV within the analysis period: Syria, Lebanon, Haiti, Dominican Republic, Iraq, Iran, and Lao People's Democratic Republic. DALY, disability-adjusted life year; NIP, national immunization programme; OCV, oral cholera vaccine; TCV, typhoid conjugate vaccine.

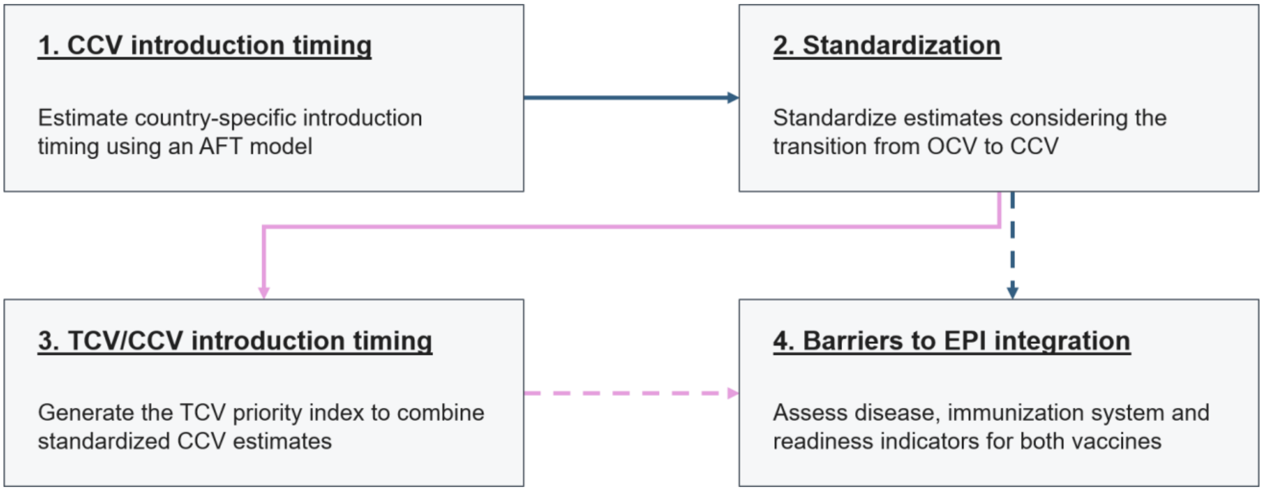


**Supplementary Figure S1. Modeling framework.** Solid arrows indicate the forecasting sequence. Dashed arrows link the vaccine pathways to the separate barrier assessment. AFT, accelerated failure time; EPI, Expanded Programme on Immunization.

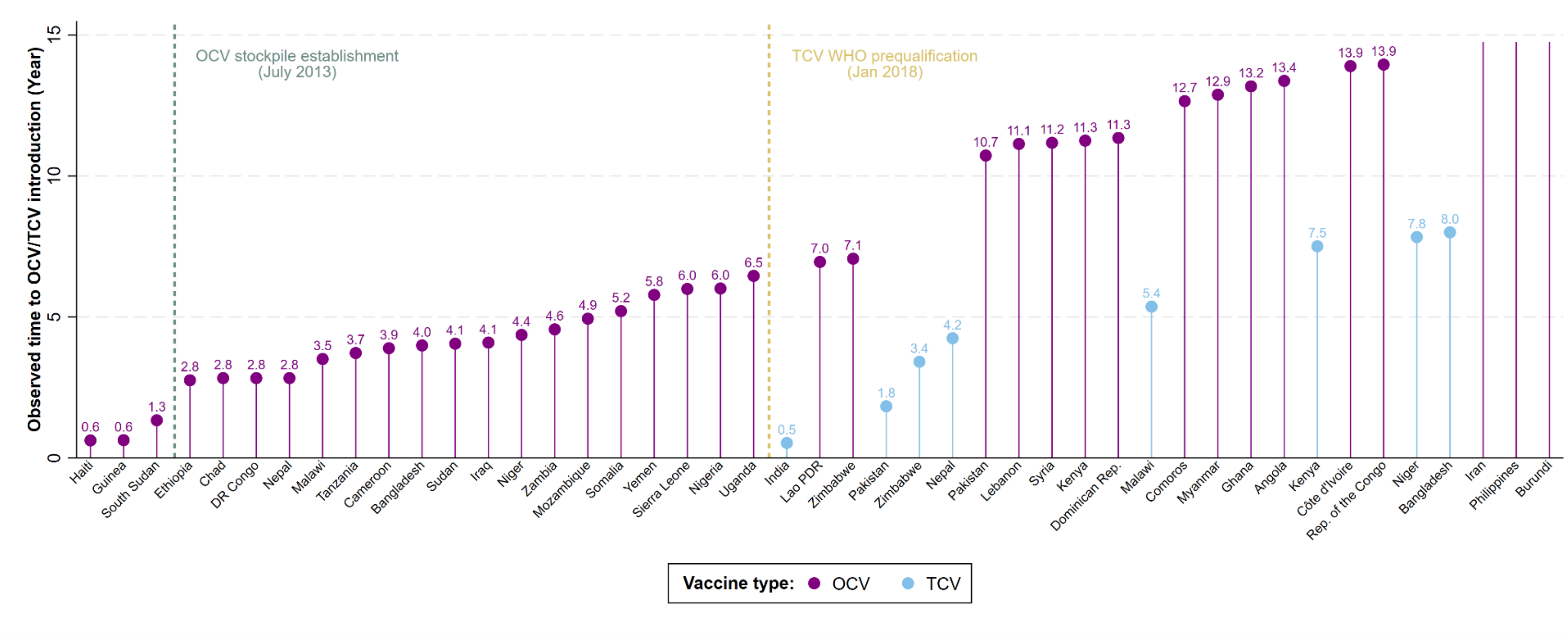


**Supplementary Figure S2. Empirical OCV, TCV introduction years.** The Philippines, Burundi, and Iran submitted OCV stockpile requests, but their requests were not approved. DR Congo, Democratic Republic of the Congo; Rep. of the Congo, Republic of the Congo; Dominican Rep., Dominican Republic; Lao PDR, Lao People's Democratic Republic.

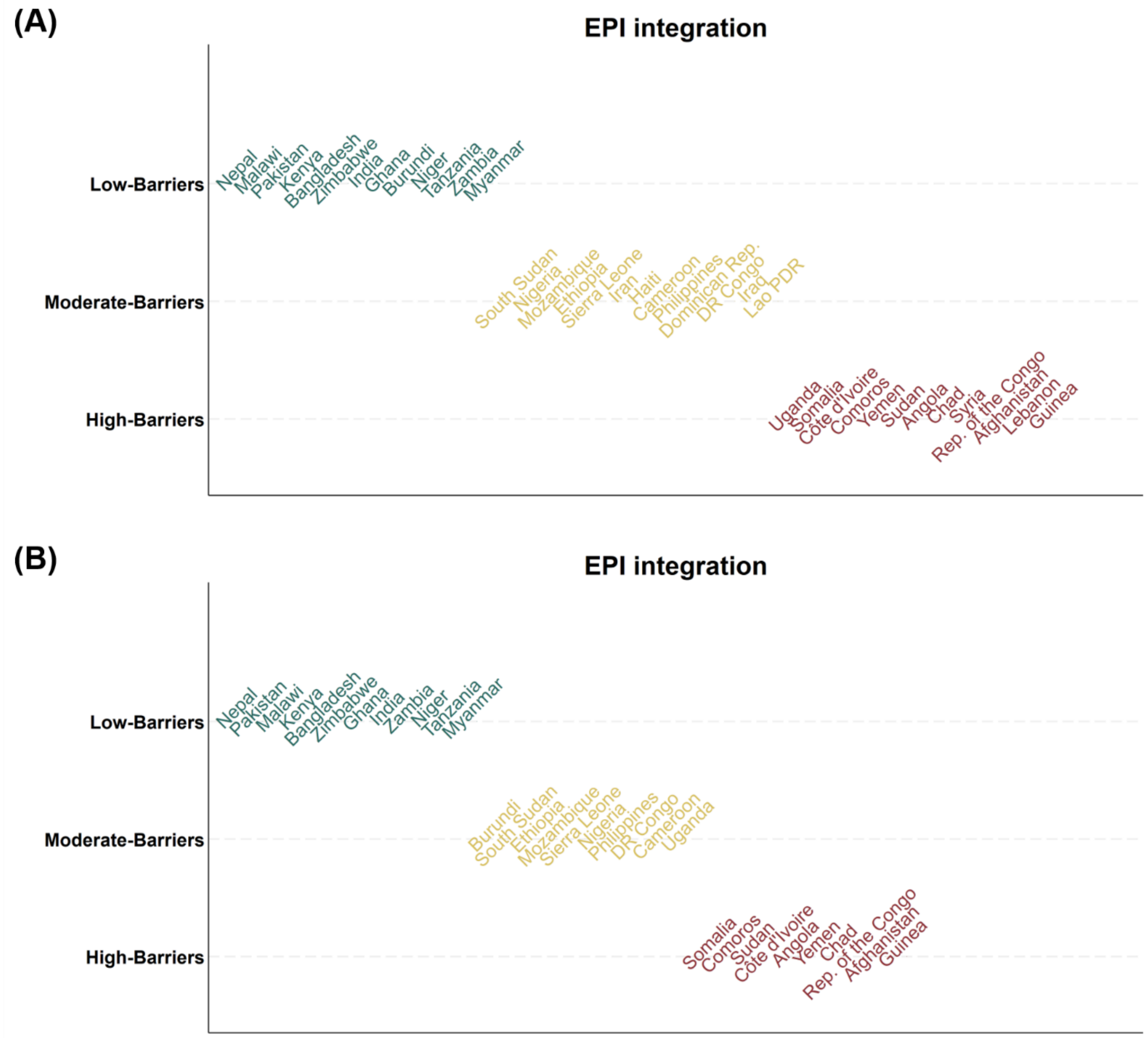


**Supplementary Figure S3. Feasibility of (A) CCV and (B) TCV/CCV into EPI.** EPI, Expanded Programme on Immunization; DR Congo, Democratic Republic of the Congo; Rep. of the Congo, Republic of the Congo; Dominican Rep., Dominican Republic; Lao PDR, Lao People's Democratic Republic.
